# Machine Learning Prediction of Autism Spectrum Disorder Through Linking Mothers’ and Children’s Electronic Health Record Data

**DOI:** 10.1101/2024.03.24.24304813

**Authors:** Yongqiu Li, Yu Huang, Shuang Yang, Elahe M. Shychuk, Elizabeth A. Shenkman, Jiang Bian, Amber M. Angell, Yi Guo

**Affiliations:** Department of Health Outcomes and Biomedical Informatics, University of Florida, Gainesville, Florida, USA; Division of Occupational Science and Occupational Therapy, University of Southern California, Los Angeles, California, USA

## Abstract

Autism spectrum disorder (ASD) is a neurodevelopmental disorder typically diagnosed in children. Early detection of ASD, particularly in girls who are often diagnosed late, can aid long-term development for children. We aimed to develop machine learning models for predicting ASD diagnosis in children, both boys and girls, using child-mother linked electronic health records (EHRs) data from a large clinical research network. Model features were children and mothers’ risk factors in EHRs, including maternal health factors. We tested XGBoost and logistic regression with Random Oversampling (ROS) and Random Undersampling (RUS) to address imbalanced data. Logistic regression with RUS considering a three-year observation window for children’s risk factors achieved the best performance for predicting ASD among the overall study population (AUROC = 0.798), boys (AUROC = 0.786), and girls (AUROC = 0.791). We calculated SHAP values to quantify the impacts of important clinical and sociodemographic risk factors.

## Introduction

Autism spectrum disorder (ASD) is a neurodevelopmental disorder typically diagnosed in children. ASD presents challenges in social interaction, and is often accompanied by repetitive behaviors and limited interests^1^. Children with ASD may also learn, move, or focus in different ways^1^. It is reported that the prevalence of ASD is 2.76% among children in the United States^2^, and the lifetime costs for supporting an individual with an ASD range from $1.4 to $2.4 million per individual^3,4^. It is crucial to identify ASD as early as possible since early intervention for ASD is associated with improved social communication, cognition, and adaptive functioning^3^. However, detecting ASD can be difficult because no medical tests, such as a blood test, exist for ASD. To screen and diagnose ASD, children are usually assessed for early sign^5^, and if of high risk, referred for clinical diagnostic evaluation^6^. Yet, no single screening or diagnostic tool is appropriate for all clinical setting, clinicians need to look at child’s developmental history and behavior to make a final diagnosis^6,7^.

Although the early reliable signs of ASD may be observed as young as two years of age, the average age of ASD diagnosis is around four years^8^. This large time gap, or delay in ASD diagnosis, may sabotage the efficacy of early interventions^7^. Furthermore, under-diagnosis of ASD is a severe problem in the United States. In the Autism and Developmental Disabilities Monitoring (ADDM) Network, the largest tracking system of ASD among children aged eight years in the United States, only 74% of the ASD children had a recorded clinical diagnosis^7^. In other words, one in four 8-year-old American children with ASD were not diagnosed, suggesting a significant under-diagnosis and highlighting the potential for late diagnosis to exacerbate psychological distress and functional challenges in daily life. The issue of delayed and under-identification of ASD is most acute among girls. Despite similar ages of parental initial concerns, the most recent surveillance data showed that the prevalence of ASD remains significantly lower among girls than among males, and research consistently finds that a critical reason for the low prevalence is under-ascertainment rather than biological or genetic causes^9^. Girls are more likely to be diagnosed later than boys, even though they do not differ from boys in the age at their parents’ “first concerns” about their development^9^.

Currently, there exist several ASD screening tools, such as the Modified Checklist for Autism in Toddlers (MCHAT). But these tools are survey instruments that require extra efforts and resources to use, and are of sub-optimal accuracy^10^. For example, despite MCHAT’s popularity as a screening tool for ASD, using it alone may not yield sufficient accuracy in detecting ASD cases. A recent analysis has demonstrated that MCHAT has a sensitivity of just 39% and a positive predictive value (PPV) of 15% for ASD detection^5^. Moreover, these instruments often fail to perform equally across population subgroups, especially in girls^11^, leading to diagnostic disparities. On the other hand, in seeking more affordable and reliable methods, electronic health record (EHR) data may be an alternative approach for early ASD detection^12^. Many risk factors for ASD can be found in EHRs, including low birth weight^13^, preterm birth^14^, low Apgar scores, other perinatal complications^14,15^ and related conditions such as attention deficit hyperactivity disorders (ADHD)^16^. While EHR data have been instrumental in the early identification of phenotypes for chronic diseases such as heart failure, diabetes, and Alzheimer’s disease before symptoms appear, using EHRs for early ASD detection is not yet fully realized. To date, EHRs have been utilized to distinguish ASD subtypes and assess suicide risk among adolescents with the condition^17^. However, there is a scarcity of studies that have used EHR data to predict ASD in children for early detection. Moreover, there is a notable lack of research that considers risk factors from both the child and the mother using EHRs, which is crucial for more accurate ASD detection^18^.

Therefore, to avoid delayed diagnosis and under-identification of ASD in children, it is imperative to build prediction models that can be easily adopted in EHRs for early and accurate ASD detection. In this study, we aimed to (1) develop gender-specific machine learning models for ASD risk prediction utilizing EHRs in the OneFlorida+ Clinical Research Consortium, part of the national Patient-Centered Clinical Research Network (PCORnet), and (2) assess the gender disparities in ASD risk prediction and risk factors by exploring feature importance in each population group.

## Methods

### Data Source and Study Population

We obtained 2012-2023 EHR data from the OneFlorida+ Clinical Research Consortium which contains patient-level information including demographics, diagnoses, medications, procedures, vital signs, lab tests, and more from 17 million residents in Florida, 2.1 million in Georgia, and 1.1 million in Alabama^19^. Our study population included a cohort of ASD children and a matching cohort of non-ASD children identified in OneFlorida+ EHRs as outlined in Figure 1.

**Figure 1.**
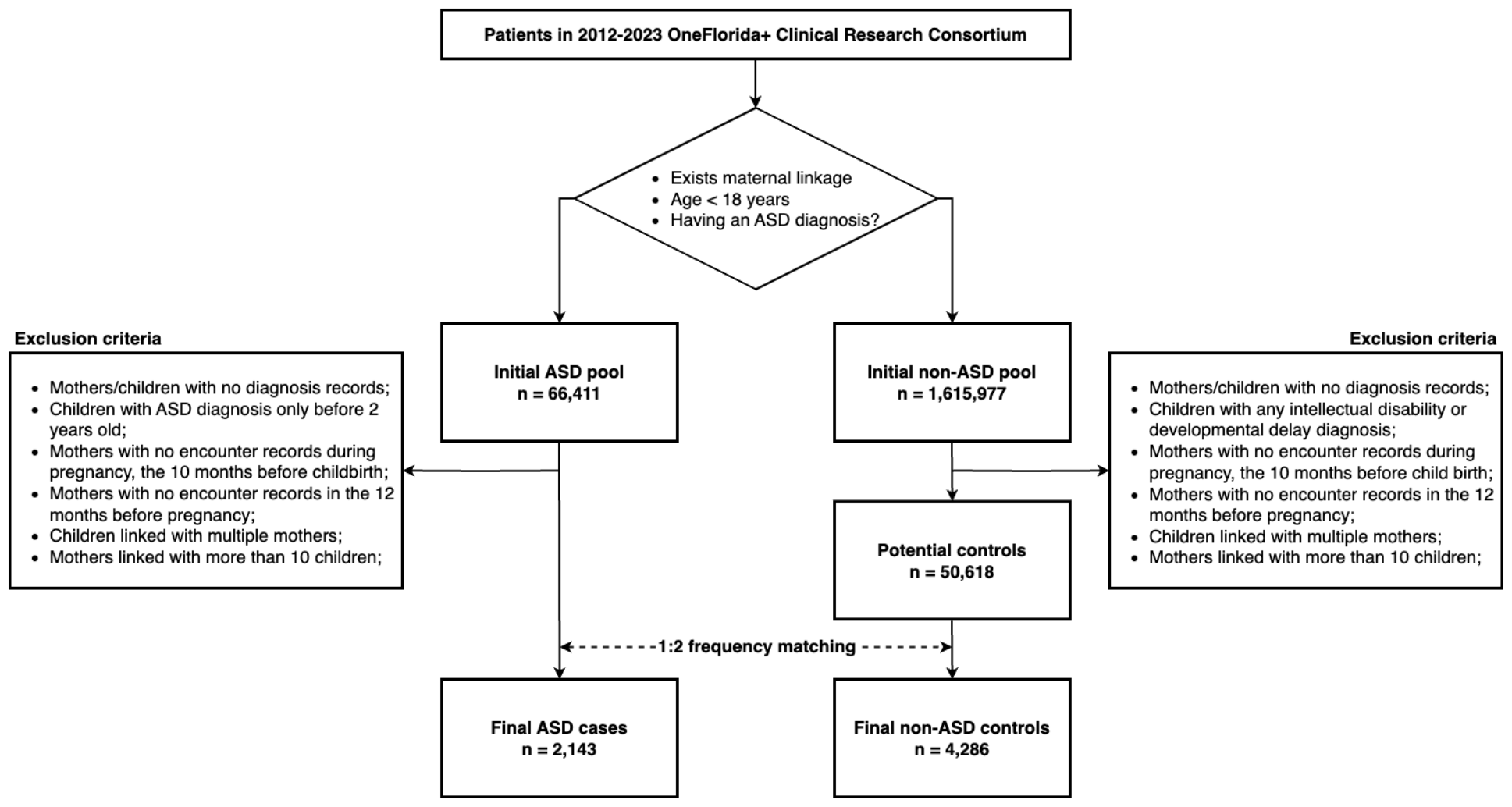
Overview of study population extraction from the OneFlorida+ clinical research consortium.

We first identified children (age < 18 years) with valid birth certificates data and therefore could be linked to their mothers’ EHR to create the initial pools of ASD and non-ASD children. ASD included Autism, Asperger’s Syndrome, and Pervasive developmental disorder not otherwise specified (PDD-NOS^7^), and was determined using International Classification of Diseases (ICD) codes (ICD-9: 299.0, 299.80, 299.9; ICD-10: F84.0, F84.5, F84.9). In the initial pools of ASD and non-ASD children with linked mother’s EHRs, we excluded: (1) mothers or children with no diagnosis records, (2) mothers with no encounter records during pregnancy (i.e., the 10 months before childbirth), (3) mothers with no encounter records in the 12 months before pregnancy, (4) children linked with multiple mothers, and (5) mothers linked with more than 10 children. For the ASD cohort, we additionally removed children with ASD diagnosis before two years old only. For the non-ASD cohort, we additionally removed children with any intellectual disability (ID) or developmental delay (DD) diagnosis. We defined the index date as the first encounter with an ASD diagnosis at or after two years old for the cases, and as a random encounter date within the index year of the case for the controls. We then matched each ASD child (i.e., cases) with two non-ASD children (i.e., controls) based on age and year.

### Overview of Data Analysis Plan

The main goal of our data analysis is to predict ASD diagnosis (ASD vs. non-ASD) using children and mothers’ risk factors in EHRs, including maternal health factors. We summarized the steps in our data analysis plan in Figure 2. In Step 1, we identified potential ASD risk factors based on a literature review and expert input. In Step 2, we extracted features associated with these factors from EHRs and balanced the dataset through preprocessing techniques. In Step 3, a set of machine learning models were trained using grid search cross-validation to optimize hyperparameters. In Step 4, we evaluated model performance using various metrics, and chose the best performing models. In Step 5, we employed SHAP^20^ (SHapley Additive exPlanations), a commonly used XAI technique, to discern important features for predicting ASD risk. Python version 3.7 with the Python libraries Sciki-learn^21^, Imbalanced-learn^22^, and statsmodels^23^ was used for data processing, machine learning modeling, and SHAP analysis.

**Figure 2.**
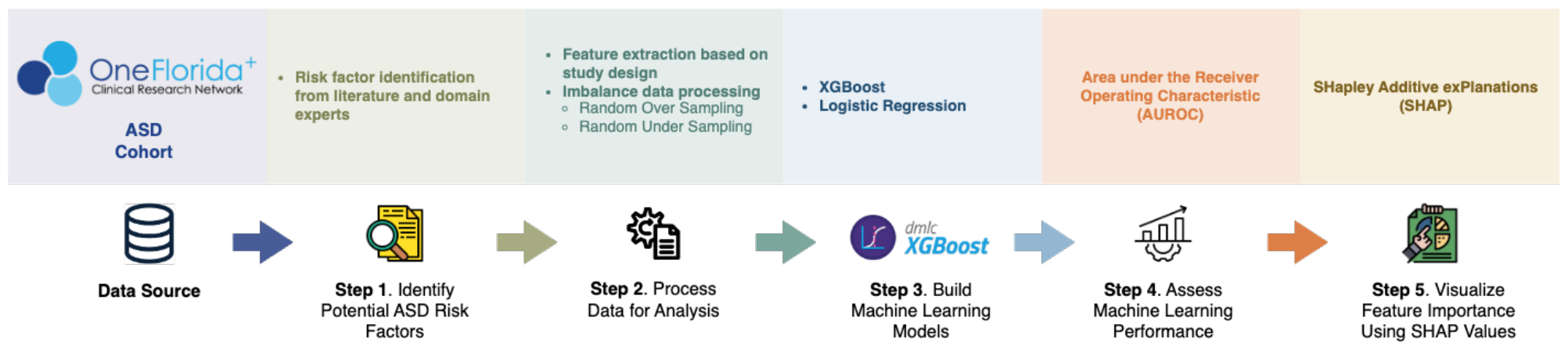
Data analysis plan overview.

#### Step 1: Identify Potential ASD Risk Factors

We conducted a review of systematic reviews from 2017–2023 by searching ASD-related terms (e.g., Autism) in PubMed to identify potential ASD risk factors associated with children and mothers (during and before pregnancy). The initial list of ASD risk factors was also reviewed and expanded by domain experts. We displayed the observation windows for risk factors associated with children and mothers in Figure 3. We adopted: (1) a varying observation window (1-, 2-, or 3-year) prior to the index date for observing risk factors from children, and (2) a 22-month observation window (10 months during pregnancy and 12 months before pregnancy) prior to childbirth for observing risk factors from mothers.

**Figure 3.**
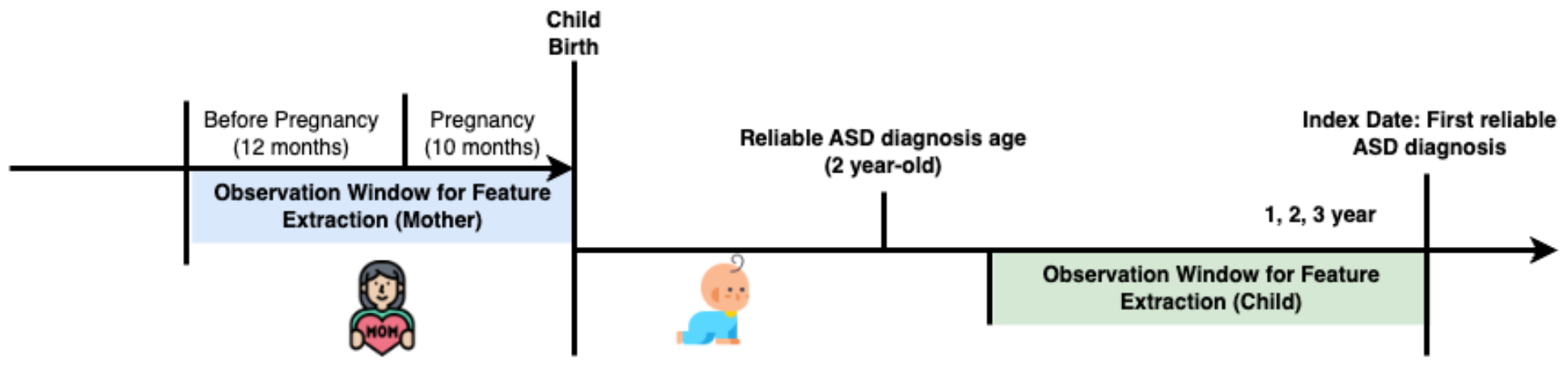
Patient timeline with observation windows for identifying risk factors.

All risk factors were identified in the children’s and their mothers’ EHRs using ICD codes, Current Procedural Terminology (CPT) codes, Healthcare Common Procedure Coding System (HCPCS) codes, RxNorm codes, and National Drug Codes (NDC). We excluded risk factors with lower than 1% prevalence within each observation window, and ultimately included 27 factors in subsequent analysis (Table 1).

**Table 1.** List of risk factors used for machine learning.

| Source | Risk Factors |
| --- | --- |
| Child | Age, Sex, Race-ethnicity, Disorders of newborn related to short gestation and low birth weight, Intellectual disabilities, Developmental delay, Congenital malformations, Genetic abnormalities, Attention-deficit/hyperactivity disorder, Anxiety |
| Mother, During pregnancy<sup>a</sup> | Age, Race-ethnicity, Diabetes, Hypertension, Psychiatric diseases, Obesity, Fever, Autoimmune diseases, Antidepressant medication |
| Mother, Pre-pregnancy<sup>b</sup> | Diabetes, Hypertension, Psychiatric diseases, Obesity, Fever, Autoimmune diseases, Abortion, Antidepressant medication |

<sup>a</sup>10 months before childbirth; <sup>b</sup>12 months before pregnancy.

#### Step 2: Process Data for Analysis

To build features for machine learning, we first coded children and mothers with any ICD, CPT, HCPCS, RxNorm and NDC code as 1’s and those without these codes as 0’s in any given observation window, since we assumed that the absence of a condition in EHR data meant the condition did not occur. We used one-hot encoding for all categorical variables. To address data imbalance (i.e., unequal distribution across outcome classes^24^), we applied random over-sampling (ROS) and random under-sampling (RUS) prior to training the models. With ROS, we duplicated data from the minority class at random (with replacement) and add them to the original dataset^24^. RUS was applied by randomly eliminating data points from the majority class, and removed them from the original dataset^24^.

#### Step 3: Build Machine Learning Models

We considered two primary classes of machine learning approaches: linear models and tree-based models. For linear models, we utilized a comprehensive set of hyperparameters and penalty functions and constructed Logistic Regression (LR) models^25^. For tree-based models, we employed Extreme Gradient Boosting (XGBoost), an algorithm renowned for its superior performance in decision-tree-based modeling, as evidenced in numerous studies^26–31^. We split the samples into training, validation, and testing sets on a ratio of 7:1:2. A five-fold cross-validation grid search was executed on the training set to optimize the model parameters. Early stopping was adopted and performed on the validation set to avoid overfitting. We developed separate models for the overall study population, girls only, and boys only. For each model, we considered three different observation windows for risk factors in children: one year, two years, or three years prior to the index date.

#### Step 4: Assess Machine Learning Performance

The performance of each machine learning model was evaluated by the area under the receiver-operating characteristic curve (AUROC), sensitivity, and specificity. We conducted bootstrapping with 100 iterations on the testing set to ensure robust performance estimates, and accounted for potential variability in the results by calculating 95% confidence intervals (CIs) for each evaluation metric^32^. Based on the AUROC, we selected the best performing model for each study population group (overall, boys and girls), resulting in a total of three models for Step 5.

#### Step 5: Visualize Feature Importance Using SHAP Values

We utilized SHAP^20^, a commonly used XAI technique, to identify important features contributing to the prediction of ASD risk and quantify their impacts. We created SHAP bar plots and summary plots highlighting the top 15 risk factors in each of the best performing models. Positive SHAP values indicated increased ASD risk, while negative values suggested decreased risk. The absolute SHAP value of each feature indicated its importance.

## Results

### Characteristics of Study Population

We summarized the basic demographics of the ASD (n = 2,143) and non-ASD (n = 4,286) cohorts in Table 2. The ASD and non-ASD children had the same age distribution (mean = 3.9 years, standard deviation = 1.8 years). There was a significantly lower percentage of girls in the ASD cohort compared to the non-ASD cohort (24.2% vs. 31.2%; p < 0.001). We observed a significant difference in the racial-ethnic distribution between the ASD and non-ASD cohorts (p < 0.001). Compared to the non-ASD cohort, the ASD cohort had lower percentages of NHW (32.0% vs. 37.4%) and NHB (24.5% vs. 25.2%) yet higher percentages of NHO (11.5% vs. 9.9%) and Hispanics (24.5% vs. 20.8%). The mothers of the ASD and non-ASD children had a similar age distribution (32.5 vs. 32.0 years, p = 0.165). Compared to the mothers of the non-ASD cohort, mothers of the ASD cohort had lower percentages of NHW (37.8% vs. 42.6%), NHB (27.4% vs. 28.1%), and NHO (6.9% vs. 7.6%), yet higher percentages of Hispanics (24.9% vs. 16.7%).

**Table 2.** Demographics of study population.

|  | ASD (n = 2,143) | Non-ASD (n = 4,286) | p-value |
| --- | --- | --- | --- |
| **Children** |  |  |  |
| Age at index date (mean, SD) | 3.9, 1.8 | 3.9, 1.8 | = 1.000 |
| Sex |  |  | < 0.001 |
| Male | 1,624 (75.8%) | 2,947 (68.8%) |  |
| Female | 519 (24.2%) | 1,339 (31.2%) |  |
| Race-ethnicity |  |  | < 0.001 |
| NHW | 686 (32.0%) | 1,604 (37.4%) |  |
| NHB | 524 (24.5%) | 1,081 (25.2%) |  |
| NHO | 247 (11.5%) | 422 (9.9%) |  |
| Hispanic | 525 (24.5%) | 891 (20.8%) |  |
| Unknown | 161 (7.5%) | 288 (6.7%) |  |
| **Mothers** |  |  |  |
| Age at childbirth (mean, SD) | 32.5, 13.1 | 32.0, 12.6 | = 0.165 |
| Race-ethnicity |  |  | < 0.001 |
| NHW | 811 (37.8%) | 1,826 (42.6%) |  |
| NHB | 587 (27.4%) | 1,205 (28.1%) |  |
| NHO | 147 (6.9%) | 327 (7.6%) |  |
| Hispanic | 534 (24.9%) | 717 (16.7%) |  |
| Unknown | 64 (3.0%) | 211 (4.9%) |  |

ASD: Autism spectrum disorder; SD: Autism spectrum disorder; NHW: non-Hispanic White; NHB: non-Hispanic Black; NHO: non-Hispanic Other.

### Performance of Machine Learning Models

We summarized the performance (i.e., AUROC, sensitivity, and specificity) of machine learning models by study population group in Table 3. Based on the AUROC, the results illustrated that the three-year observation window was the optimal period for ASD prediction across all study population groups as compared to the one- or two-year observation window in most cases. We also found that RUS consistently produced satisfactory results across different study population groups and observation windows, and logistic regression consistently outperformed XGboost, although slightly. Under RUS, logistic regression with a three-year observation window achieved the best AUROC of 0.786, 0.791, 0.798 for predicting ASD in the overall, boys, and girls study population, respectively.

**Table 3.** Overall performance of machine models.

|  |  | AUROC |  |  | Sensitivity |  |  | Specificity |  |  |
| --- | --- | --- | --- | --- | --- | --- | --- | --- | --- | --- |
|  |  | 1-year | 2-year | 3-year | 1-year | 2-year | 3-year | 1-year | 2-year | 3-year |
| **Overall (Boys + Girls)** |  |  |  |  |  |  |  |  |  |  |
| ROS | LR | 0.789 | 0.795 | 0.798 | 0.576 | 0.584 | 0.588 | 0.886 | 0.887 | 0.891 |
|  | XGboost | 0.785 | 0.793 | 0.796 | 0.526 | 0.558 | 0.569 | 0.921 | 0.899 | 0.899 |
| RUS | LR | 0.789 | 0.795 | 0.798 | 0.578 | 0.581 | 0.590 | 0.883 | 0.891 | 0.888 |
|  | XGboost | 0.782 | 0.790 | 0.794 | 0.525 | 0.565 | 0.544 | 0.925 | 0.895 | 0.921 |
| **Boys** |  |  |  |  |  |  |  |  |  |  |
| ROS | LR | 0.774 | 0.781 | 0.786 | 0.561 | 0.571 | 0.578 | 0.866 | 0.874 | 0.874 |
|  | XGboost | 0.765 | 0.775 | 0.781 | 0.561 | 0.514 | 0.540 | 0.866 | 0.922 | 0.897 |
| RUS | LR | 0.774 | 0.781 | 0.786 | 0.531 | 0.567 | 0.584 | 0.884 | 0.878 | 0.872 |
|  | XGboost | 0.767 | 0.769 | 0.780 | 0.566 | 0.553 | 0.520 | 0.863 | 0.871 | 0.925 |
| **Girls** |  |  |  |  |  |  |  |  |  |  |
| ROS | LR | 0.778 | 0.789 | 0.784 | 0.566 | 0.572 | 0.530 | 0.877 | 0.905 | 0.937 |
|  | XGboost | 0.776 | 0.786 | 0.784 | 0.483 | 0.514 | 0.527 | 0.971 | 0.957 | 0.952 |
| RUS | LR | 0.782 | 0.788 | 0.791 | 0.570 | 0.577 | 0.577 | 0.883 | 0.891 | 0.888 |
|  | XGboost | 0.768 | 0.773 | 0.784 | 0.494 | 0.512 | 0.544 | 0.959 | 0.956 | 0.935 |

ROS: Random Oversampling; RUS: Random Undersampling, LR: Logistic Regression.
1-year, 2-year, 3-year are varying observation windows before index for children's risk factors.

### SHAP Values Measuring Feature Importance

We calculated SHAP values based on the best performing model, logistic regression with RUS considering a three-year observation window for children’s risk factors. We displayed the SHAP summary plots for the top 15 features in each population group in Figure 4.

**Figure 4.**
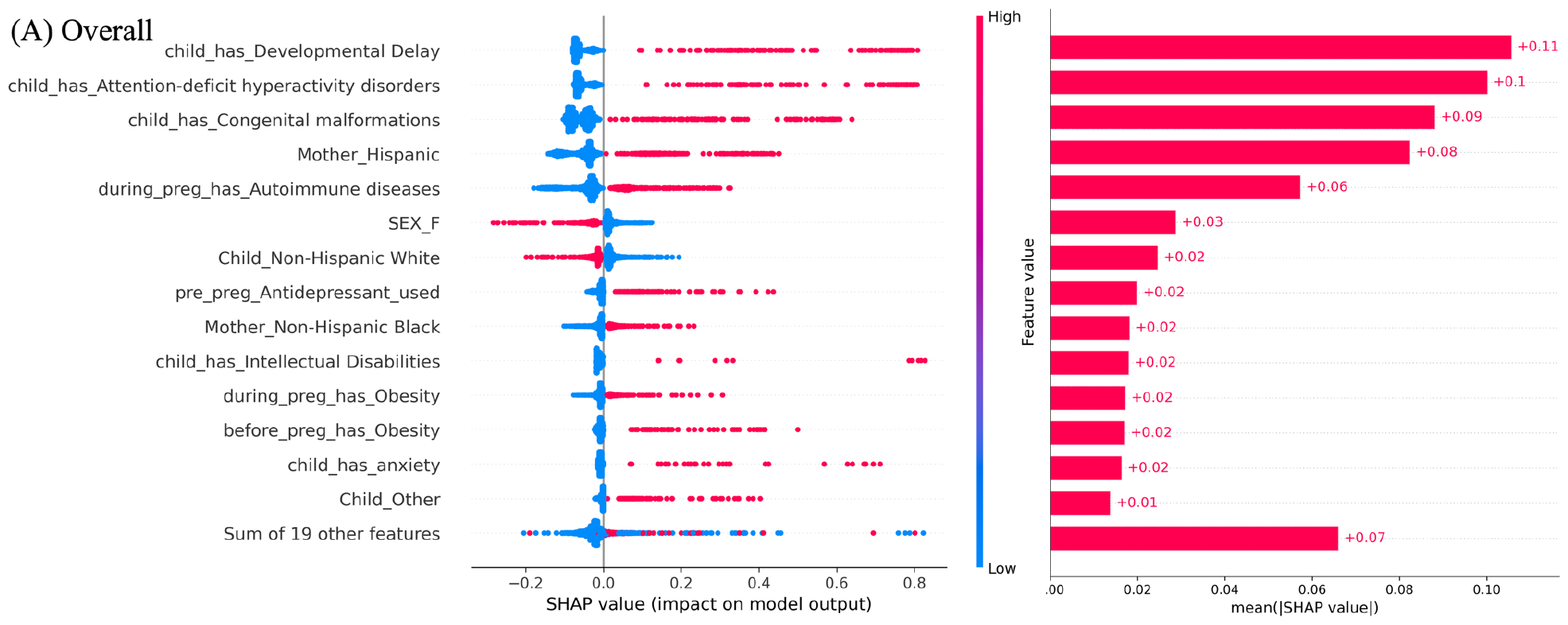
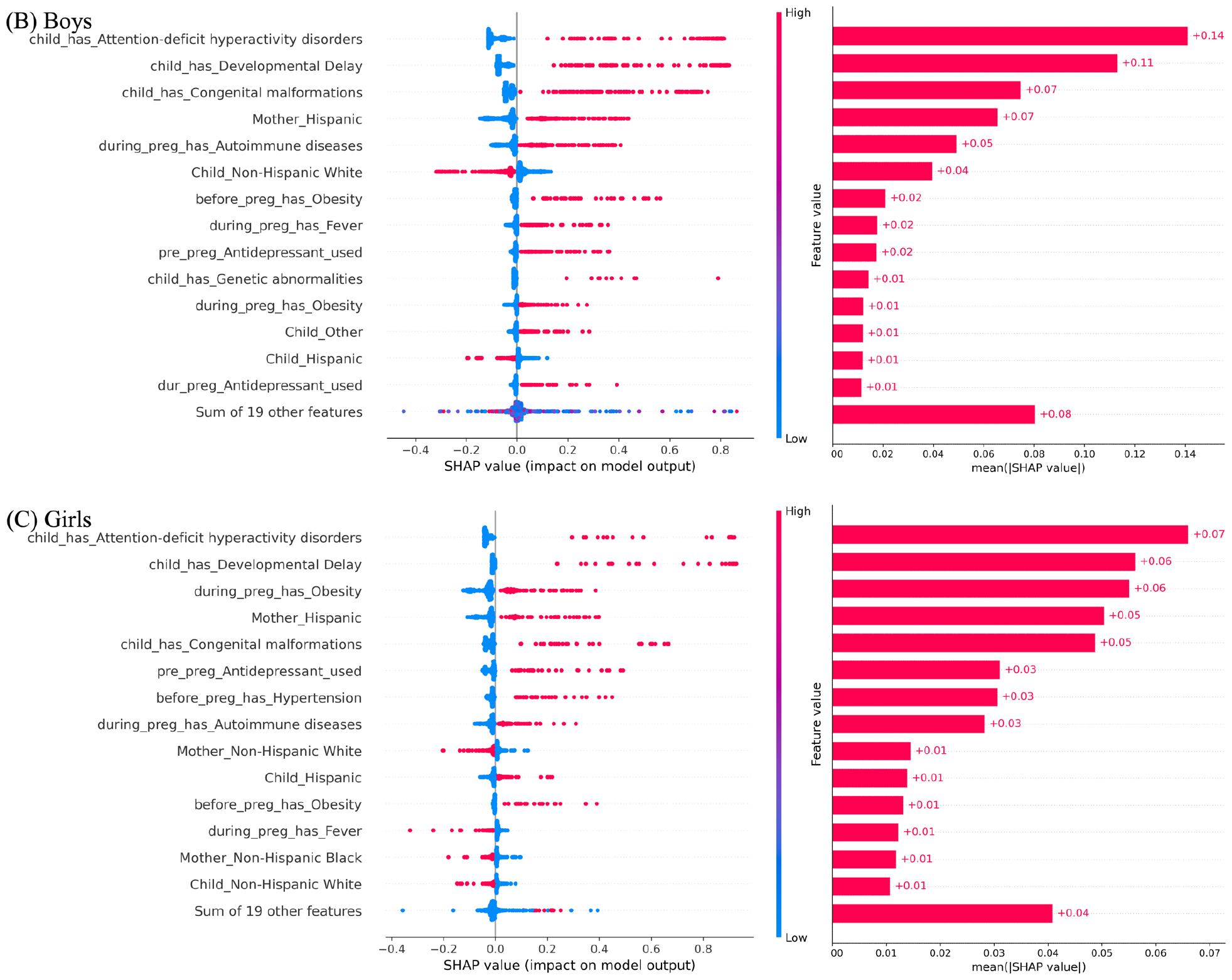
SHAP plots of the top 15 features from logistic regression using the RUS resampling technique based on a three-year observation window for children’s ASD risk factor. (A) Overall (i.e., both boys and girls), (b) boys, and (c) girls.

Figure 4A shows the important features for the overall study population of children. DD and ADHD were the two most impactful predictors for increased risk of ASD. Being NHO or having diagnoses such as congenital malformations, ID, and anxiety was associated with elevated ASD risk, whereas being girl or NHW was associated with lower risk of ASD. Additionally, risk factors from the mothers also played a significant role in ASD risk prediction. Significant maternal health risk factors for ASD included having autoimmune diseases and obesity. Mother being Hispanic or NHB as well as pre-pregnancy obesity and use of antidepressant medications were all associated with increased risk of ASD.

Figure 4B highlights the influential factors for predicting ASD in boys. ADHD and DD were marked as the two most significant predictors of ASD. Other predictors contributing to increased risk of ASD included congenital malformations and genetic abnormalities. Being Hispanic or NHW was associated with a decreased ASD risk, whereas being NHO was associated with an increased ASD risk. Maternal health risk factors included the use of antidepressant medications and having fever, obesity, and autoimmune diseases. Other risk factors from mothers included being Hispanic as well as pre-pregnancy obesity and use of antidepressant medications.

Figure 4C highlights the important factors for predicting ASD risk in girls. The plots revealed that the diagnosis of ADHD and DD were the two primary predictors of ASD risk. Other predictors such as congenital malformations also contributed to an increased risk of ASD. Being Hispanic was associated with an increased ASD risk, while being NHW was associated with a decreased ASD risk. Maternal health risk factors for ASD such as having autoimmune diseases and obesity were associated an increased ASD risk, whereas fever was associated with a decreased ASD risk. Regarding other risk factors from mothers, being Hispanic as well as having pre-pregnancy hypertension, obesity and use of antidepressant medications were associated with an increased risk of ASD, while being NHB or NHW was associated with a decreased risk of ASD.

## Discussion and Conclusions

In this study, we developed machine learning models for identifying children with ASD within three study population groups, including overall (i.e., both boys and girls), boys, and girls using OneFlorida+ EHR data. We tested both XGBoost and logistic regression using the ROS and RUS resampling techniques to create the prediction models considering a varying observation window (i.e., one-year, two-year, or three-year before index date) for children’s ASD risk factors. Logistic regression using the RUS resampling method with a three-year observation window achieved the best performance for predicting ASD among the overall population (AUROC = 0.798), boys (AUROC = 0.786), and girls (AUROC = 0.791), which indicates that our final prediction models are accurate tools for predicting ASD among children. Particularly, the model for girls achieved an AUROC of 0.791, which is as good as the model for boys and the overall population, indicating our model can be used to detect ASD in girls who are usually face delays in diagnosis.

In each population group (i.e., overall, boys, and girls), we identified important features for ASD risk prediction by calculating SHAP values using our three best performing models. We observed some of the same predictors in the top features in all population groups. In particular, DD and ADHD were consistently underscored as the most important predictors of ASD in all groups, which aligns with existing literature. For instance, Riaz et al reported that the majority of children with ASD experience did not achieve developmental milestones and suffered from developmental delays in a few skills such as language and social skills^33^. Similarly, ADHD’s co-occurrence with ASD was supported by several studies, which suggested that 20-50% of children with ADHD also fulfilled the criteria for ASD, and 30-80% of children with ASD might exhibit symptoms of ADHD^34–40^. These prevalence rates serve to rationalize the prominence of these features in our models across all population groups. In the model for the overall study population (i.e., both boys and girls) (Figure 4A), we observed a reduced ASD risk in girls, which aligns with previous findings that girls often face delays and underdiagnosis for ASD^9^. Examination of the SHAP plots also highlighted that congenital abnormalities had a more pronounced impact in boys. This could be attributed to the higher prevalence of most congenital anomalies in males at birth, as supported by research from Lary et al^41^.

Maternal health factors, such as having autoimmune diseases and obesity were consistently shown as predictors of ASD in all population groups. This finding of ours aligns with existing literature. For instance, Chen et al found that maternal autoimmune diseases significantly increased the risk of ASD in children^42^. Similarly, Gardner et al revealed that maternal obesity was associated with an increased ASD risk^43^. Other factors from mothers such as being Hispanic as well as pre-pregnancy obesity and use of antidepressant medications were also showed to consistently correlate with increased ASD risk in all population groups. Andrade et al reported that antidepressant use 3-12 months prior to pregnancy was correlated with a rise in ASD risk in children^44^. Krakowlak et al observed that children whose mother was obese before pregnancy had 67% higher risk of ASD^45^. The identification of ASD risk factors from mothers, independent of those from children, underscores the value of linking children’s and mothers’ EHRs, which offers a more comprehensive view of factors contributing to ASD. This unique approach of ours indicates the importance of considering risk factors from mothers, both pre-pregnancy factors and maternal health factors, a perspective not extensively explored in prior EHR studies.

Our study has several strengths, primarily the innovative integration and analysis of linked mother-child EHR data, which provided a more comprehensive perspective on potential ASD risk factors from both maternal and child health. Furthermore, our gender-specific models, which showed good performance in both boys and girls, may be leveraged to accurately predict ASD in girls, who are typically under-diagnosed. Our study is not without limitations. Although we utilized EHR data from a large clinical research network, the models we built were not externally validated across diverse datasets, which may limit the generalizability of our findings. Additionally, we were unable to examine certain risk factors potentially important for ASD prediction, such as family history (e.g., risk factors related to father) and environmental exposures, due to the lack of data on these factors in EHRs. Future studies should aim to include these important yet omitted factors for ASD prediction, and validate the resulted models in multi-site settings to ensure the models’ applicability on a broader scale.

In summary, our study developed machine learning prediction models for ASD in the overall children population, and boys and girls separately. Our models used child-mother linked EHR data and can be used for accurate ASD prediction in both boys as well as girls who are usually under-diagnosed. Future research should prioritize external model validation and investigate the models’ clinical applications as clinical decision support tools. Incorporating additional data types, such as clinical notes, genomic data, and exposome data, could further be considered to enhance the predictive ability of ASD risk models.

## Data Availability

All data produced in the present study are available upon reasonable request to the authors

## Acknowledgement

This study was supported by grant R21MH129682 from the National Institutes of Health (NIH). This study was also partially supported by NIH grants R01CA246418, R01CA246418-02S1, R21CA245858, R21CA245858-01A1S1, R21CA253394-01A1, R01AG080624, and R21AG068717. The authors wish to thank the Cancer Informatics Shared Resource in the UF Health Cancer Center for data analytics support.

## References

1. CDC. Signs and symptoms of autism Spectrum Disorder [Homepage on the Internet]. Centers for Disease Control and Prevention. 2023 [cited 2024 Mar 16];Available from: https://www.cdc.gov/ncbddd/autism/signs.html

2. Shaw KA, Bilder DA, McArthur D, et al. Early identification of autism spectrum disorder among children aged 4 years - autism and Developmental Disabilities Monitoring Network, 11 sites, United States, 2020. MMWR Surveill Summ 2023;72(1):1–15.

3. Zwaigenbaum L, Bauman ML, Choueiri R, et al. Early intervention for children with autism spectrum disorder under 3 years of age: Recommendations for practice and research. In: Pediatric Collections: Autism Spectrum Disorder. American Academy of Pediatrics, 2020; p. 269–290.

4. Buescher AVS, Cidav Z, Knapp M, Mandell DS. Costs of autism spectrum disorders in the United Kingdom and the United States. JAMA Pediatr 2014;168(8):721.

5. Guthrie W, Wallis K, Bennett A, et al. Accuracy of autism screening in a large pediatric network. In: Pediatric Collections: Autism Spectrum Disorder. American Academy of Pediatrics, 2020; p. 101–112.

6. Identification, evaluation, and management of children with autism spectrum disorder. In: Pediatric Clinical Practice Guidelines & Policies, 21st Ed. American Academy of Pediatrics, 2021; p. 855–925.

7. CDC. Screening and diagnosis of autism spectrum disorder [Homepage on the Internet]. Centers for Disease Control and Prevention. 2022 [cited 2024 Mar 12];Available from: https://www.cdc.gov/ncbddd/autism/screening.html

8. Maenner MJ, Shaw KA, Bakian AV, et al. Prevalence and characteristics of autism spectrum disorder among children aged 8 years — autism and developmental disabilities monitoring network, 11 sites, United States, 2018. Morb Mortal Wkly Rep Surveill Summ 2021;70(11):1–16.

9. Maenner MJ, Warren Z, Williams AR, et al. Prevalence and characteristics of autism spectrum disorder among children aged 8 years — autism and developmental disabilities monitoring network, 11 sites, United States, 2020. Morb Mortal Wkly Rep Surveill Summ 2023;72(2):1–14.

10. Cavus N, Lawan AA, Ibrahim Z, et al. A systematic literature review on the application of machine-learning models in behavioral assessment of autism spectrum disorder. J Pers Med 2021;11(4):299.

11. Begeer S, Mandell D, Wijnker-Holmes B, et al. Sex differences in the timing of identification among children and adults with autism spectrum disorders. J Autism Dev Disord 2013;43(5):1151–1156.

12. Engelhard MM, Henao R, Berchuck SI, et al. Predictive value of early autism detection models based on electronic health record data collected before age 1 year. JAMA Netw Open 2023;6(2):e2254303.

13. Gardener H, Spiegelman D, Buka SL. Perinatal and neonatal risk factors for autism: A comprehensive meta-analysis. Pediatrics 2011;128(2):344–355.

14. Fassett M, Peltier M, Wing D, et al. Association of perinatal risk factors with autism spectrum disorder. Am J Perinatol 2017;34(03):295–304.

15. Buchmayer S, Johansson S, Johansson A, Hultman CM, Sparén P, Cnattingius S. Can association between preterm birth and autism be explained by maternal or neonatal morbidity? Pediatrics 2009;124(5):e817–e825.

16. Slaby I, Hain HS, Abrams D, et al. An electronic health record (EHR) phenotype algorithm to identify patients with attention deficit hyperactivity disorders (ADHD) and psychiatric comorbidities. J Neurodev Disord [homepage on the Internet] 2022;14(1). Available from: 10.1186/s11689-022-09447-9

17. Chen Y-H, Chen Q, Kong L, Liu G. Early detection of autism spectrum disorder in young children with machine learning using medical claims data. BMJ Health Care Inform 2022;29(1):e100544.

18. Ou J, Dong H, Dai S, et al. Development and validation of a risk score model for predicting autism based on pre- and perinatal factors. Front Psychiatry [homepage on the Internet] 2024 [cited 2024 Mar 12];15. Available from: https://www.ncbi.nlm.nih.gov/pmc/articles/PMC10904522/

19. OneFlorida+ – clinical research network [Homepage on the Internet]. [cited 2024 Mar 7];Available from: https://onefloridaconsortium.org/

20. Lundberg S, Lee S-I. A unified approach to interpreting model predictions [Homepage on the Internet]. arXiv [cs.AI]. 2017 [cited 2023 Jan 15];Available from: https://proceedings.neurips.cc/paper/2017/hash/8a20a8621978632d76c43dfd28b67767-Abstract.html

21. Pedregosa F, Varoquaux G, Gramfort A, et al. Scikit-learn: Machine Learning in Python. J Mach Learn Res 2011;12(85):2825–2830.

22. Lemaitre G, Nogueira F, Aridas CK. Imbalanced-learn: A python toolbox to tackle the curse of imbalanced datasets in machine learning [Homepage on the Internet]. arXiv [cs.LG]. 2016 [cited 2023 Feb 8];Available from: https://www.jmlr.org/papers/volume18/16-365/16-365.pdf

23. Seabold S, Perktold J. Statsmodels: Econometric and statistical modeling with python [Homepage on the Internet]. In: Proceedings of the 9th Python in Science Conference. SciPy, 2010 [cited 2023 Jul 19]; Available from: https://conference.scipy.org/proceedings/scipy2010/seabold.html

24. Yang C, Fridgeirsson EA, Kors JA, Reps JM, Rijnbeek PR. Impact of random oversampling and random undersampling on the performance of prediction models developed using observational health data. J Big Data [homepage on the Internet] 2024;11(1). Available from: 10.1186/s40537-023-00857-7

25. Tolles J, Meurer WJ. Logistic Regression: Relating Patient Characteristics to Outcomes. JAMA 2016;316(5):533–534.

26. Shin J, Lee J, Ko T, Lee K, Choi Y, Kim H-S. Improving Machine Learning Diabetes Prediction Models for the Utmost Clinical Effectiveness. J Pers Med [homepage on the Internet] 2022;12(11). Available from: 10.3390/jpm12111899

27. Zhao Y, Li X, Li S, et al. Using Machine Learning Techniques to Develop Risk Prediction Models for the Risk of Incident Diabetic Retinopathy Among Patients With Type 2 Diabetes Mellitus: A Cohort Study. Front Endocrinol 2022;13:876559.

28. Deberneh HM, Kim I. Prediction of Type 2 Diabetes Based on Machine Learning Algorithm. Int J Environ Res Public Health [homepage on the Internet] 2021;18(6). Available from: 10.3390/ijerph18063317

29. Li Y, Wang H, Luo Y. Improving Fairness in the Prediction of Heart Failure Length of Stay and Mortality by Integrating Social Determinants of Health. Circ Heart Fail 2022;15(11):e009473.

30. Yang H, Li J, Liu S, Yang X, Liu J. Predicting Risk of Hypoglycemia in Patients With Type 2 Diabetes by Electronic Health Record-Based Machine Learning: Development and Validation. JMIR Med Inform 2022;10(6):e36958.

31. Wang L, Wang X, Chen A, Jin X, Che H. Prediction of Type 2 Diabetes Risk and Its Effect Evaluation Based on the XGBoost Model. Healthcare (Basel) [homepage on the Internet] 2020;8(3). Available from: 10.3390/healthcare8030247

32. Efron B, Tibshirani R. Correction to: The bootstrap method for assessing statistical accuracy. Behaviormetrika 2021;48(1):191–191.

33. Patterns of Developmental Delay in Children with Autism Spectrum Disorder: A Perspective from a Developing Country.

34. Hattori J, Ogino T, Abiru K, Nakano K, Oka M, Ohtsuka Y. Are pervasive developmental disorders and attention-deficit/hyperactivity disorder distinct disorders? Brain Dev 2006;28(6):371–374.

35. Frazier JA, Biederman J, Bellordre CA, et al. Should the diagnosis of Attention-Deficit/ Hyperactivity disorder be considered in children with Pervasive Developmental Disorder? J Atten Disord 2001;4(4):203–211.

36. Holtmann M, Bölte S, Poustka F. Attention deficit hyperactivity disorder symptoms in pervasive developmental disorders: Association with autistic behavior domains and coexisting psychopathology. Psychopathology 2007;40(3):172–177.

37. Lee DO, Ousley OY. Attention-deficit hyperactivity disorder symptoms in a clinic sample of children and adolescents with pervasive developmental disorders. J Child Adolesc Psychopharmacol 2006;16(6):737–746.

38. Rowlandson PH, Smith C. An interagency service delivery model for autistic spectrum disorders and attention deficit hyperactivity disorder. Child Care Health Dev 2009;35(5):681–690.

39. Simonoff E, Pickles A, Charman T, Chandler S, Loucas T, Baird G. Psychiatric disorders in children with autism spectrum disorders: prevalence, comorbidity, and associated factors in a population-derived sample. J Am Acad Child Adolesc Psychiatry 2008;47(8):921–929.

40. Yoshida Y, Uchiyama T. The clinical necessity for assessing Attention Deficit/Hyperactivity Disorder (AD/HD) symptoms in children with high-functioning Pervasive Developmental Disorder (PDD). Eur Child Adolesc Psychiatry 2004;13(5):307–314.

41. Lary JM, Paulozzi LJ. Sex differences in the prevalence of human birth defects: A population-based study. Teratology 2001;64(5):237–251.

42. Chen C-C, Lin C-H, Lin M-C. Maternal autoimmune disease and risk of offspring autism spectrum disorder – a nationwide population-based cohort study. Front Psychiatry [homepage on the Internet] 2023;14. Available from: 10.3389/fpsyt.2023.1254453

43. Gardner RM, Lee BK, Magnusson C, et al. Maternal body mass index during early pregnancy, gestational weight gain, and risk of autism spectrum disorders: Results from a Swedish total population and discordant sibling study. Int J Epidemiol 2015;44(3):870–883.

44. Andrade C. Antidepressant exposure during pregnancy and risk of autism in the offspring, 2: Do the new studies add anything new? J Clin Psychiatry 2017;78(8):e1052–e1056.

45. Krakowiak P, Walker CK, Bremer AA, et al. Maternal metabolic conditions and risk for autism and other neurodevelopmental disorders. Pediatrics 2012;129(5):e1121–8.

